# Diagnosis of SARS-CoV-2 infections in times of material shortage

**DOI:** 10.1101/2020.12.01.20242008

**Authors:** Thaisa Lucas Sandri, Juliana Inoue, Johanna Geiger, Johanna-Marie Griesbaum, Constanze Heizel, Michael Burnet, Rolf Fendel, Peter G. Kremsner, Jana Held, Andrea Kreidenweiss

## Abstract

The pandemic caused by SARS-CoV-2 resulted in increasing demands for diagnostic tests, leading to a shortage of recommended testing materials and reagents. This study reports on the performance of self-sampled alternative swabbing material (ordinary Q-tips tested against flocked swab and rayon swab), of reagents for classical RNA extraction (phenol/guanidine-based protocol against a commercial kit), and of intercalating dye-based one-step quantitative reverse transcription real-time PCRs (RT-qPCR) compared against the gold standard hydrolysis probe-based assays for SARS-CoV-2 detection. The study found sampling with Q-tips, RNA extraction with classical protocol and intercalating dye-based RT-qPCR as a reliable and comparably sensitive strategy for detection of SARS-CoV-2 - particularly valuable in the current period with a resurgent and dramatic increase in SARS-CoV-2 infections and growing shortage of diagnostic materials as well for regions limited in resources.

## Introduction

At the end of 2019, the new coronavirus SARS-CoV-2 (severe acute respiratory syndrome coronavirus 2) was discovered in Wuhan, China, causing the respiratory disease COVID-19. The following outbreak, classified by the World Health Organisation (WHO) as a pandemic situation on 11^th^ March 2020, resulted in increasing demand for fast and reliable diagnostic tests to identify infected individuals to prevent the spread of the virus (1, 2). Many health institutions reported shortages of required materials for pharyngeal specimen collection, sample extraction, and PCR materials. To ensure reliable diagnosis of SARS-CoV-2 infections in the event of a resurgence in infection rates, such as those currently observed in Europe and North America in autumn/winter 2020/2021, suitable alternative materials need to be urgently identified (1, 2).

Although rapid tests for SARS-CoV-2 antigen detection are becoming available, reverse transcription quantitative real-time PCR (RT-qPCR) will remain the gold standard for diagnosing SARS-CoV-2 infections due to excellent sensitivity. Until very recently, there were only few approved materials, reagents, and procedures for SARS-CoV-2 molecular diagnostics (3–5). Guidelines recommended the preferred use of flocked swabs stored in transport medium over dry swabs collected by specialists (6) and listed only kit-based RNA extraction methodologies followed by a hydrolysis probe-based RT-qPCR assay. These materials and methods became the gold standard procedure for detection of SARS-CoV-2.

With the increasing demand for SARS-CoV-2 diagnostic tests for people suspected of having COVID-19 and population screenings, there is a continuing shortage of recommended swabs, RNA extraction kits, and reagents for hydrolysis probe-based RT-qPCR. In addition, the difficulty of providing rapid access and the high costs of recommended consumables pose a barrier for wider application of this diagnostic method for low-resource countries. Therefore, adaptation of diagnostic procedures to available and alternative reagents is necessary to ensure continued reliable diagnosis. For this purpose, we tested self-sampled, ordinary cotton swabs “Q-tips” as an alternative to medical swabs for oropharyngeal sampling, the classical RNA extraction protocol, and a cost-saving intercalating dye-based RT-qPCR assay for the detection of SARS-CoV-2.

## Materials and Methods

### Study participants

A total of 13 participants were enrolled in the study, seven individuals with acute SARS-CoV-2 infection and six uninfected control individuals. Study participants were requested to sample themselves and the procedure of oropharyngeal swabbing was explained to each participant. Each participant was provided with two kits containing three different swab types each that were collected by the study team within two hours after sampling. All individuals gave written informed consent. The study was approved by the Ethics Committee of the Universitätsklinikum Tübingen (Ref. number 20/231/B01).

### Sample collection

Three different swab types were compared for their applicability to oropharyngeal sampling and detection of SARS-CoV-2 infections: 1) Q-tips, these are ordinary cotton swabs with a paper stick usually used for ear cleaning and available in local supermarkets, 2) flocked swab embedded in 2 ml of 0.9% NaCl solution after sampling (COPAN FLOQswab 501CS01, Copan, Italy), and 3) dry rayon swab (COPAN 155C Rayon, Copan, Italy). Swabs were kept at 4°C until RNA extraction but no longer than 4 hours.

### RNA extraction

Two different RNA extraction methodologies were compared for their efficiency in RNA yield: a classical protocol using phenol/guanidine thiocyanate (QIAzol Lysis Reagent, QIAGEN) for cell lysis and RNase inhibition, chloroform and isopropanol for RNA purification and precipitation, respectively, following manufacturer’s protocol (7), and the commercial kit QIAamp Viral RNA Mini kit (QIAGEN). To start the RNA extraction, the complete cotton material of Q-tips and rayon swabs was cut and either immersed in 1 ml QIAzol or 560 µl AVL buffer containing carrier RNA when utilizing QIAamp Viral RNA Mini kit and kept for 10 minutes at room temperature before continuing with the respective protocols. Flocked swabs were stored in 2 ml saline after sampling, 140 µl of this solution was used for RNA extraction for each one of the two extraction methodologies. Total RNA was eluted with 60 µl nuclease-free water for the classical extraction or elution buffer provided by the kit. RNA concentration and quality per extraction were measured using NanoDrop 1000 (NanoDrop Technologies).

### Endogenous human control

Detection of human RNAse P (hRNAse P) RNA in swab material can serve as an endogenous control, to ensure sample integrity and proper RNA extraction as suggested by the CDC (5). Here, we used hRNAse P one-step reverse transcription quantitative real-time PCR (RT-qPCR) (**Table 1**) to ensure proper sampling and RNA extraction when using different swab types and to evaluate RNA extraction protocols using samples from SARS-CoV-2 non-infected participants. Synthetic hRNAse P RNA was used for the standard curves and as positive control.

**Table 1.**
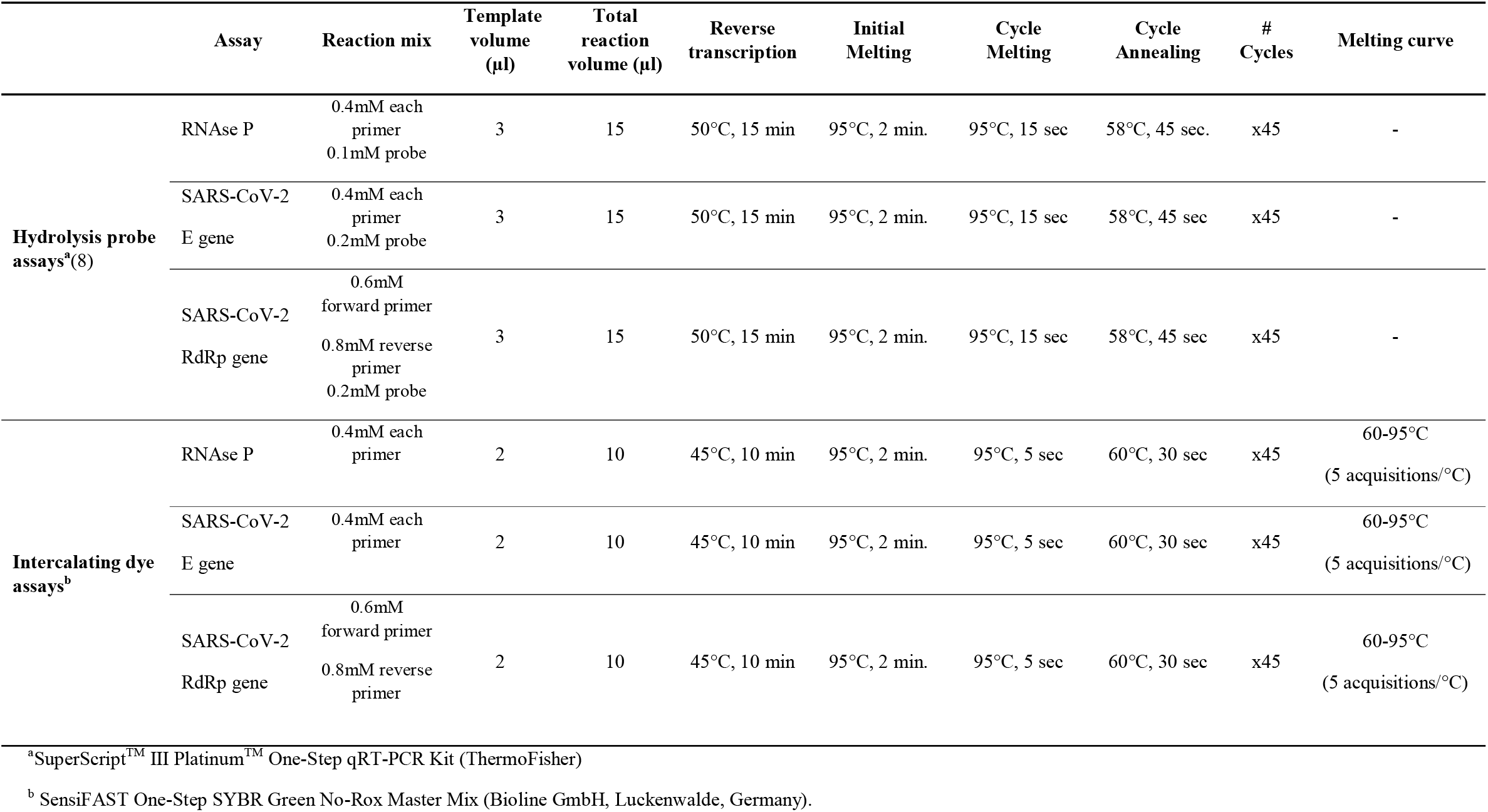
Nucleic acid amplification assays conditions

### SARS-CoV-2 assays

For SARS-CoV-2 detection, the protocols for hydrolysis probe-based, one-step, reverse transcription quantitative real-time PCR (probe RT-qPCR) from the Institute of Virology, Charité, Berlin, Germany, targeting the envelope (E) and the RNA dependent RNA polymerase (RdRp) genes of SARS-CoV-2 were utilized (8). These protocols were further modified to establish intercalating dye-based, one-step, reverse transcription quantitative real-time PCR (SYBR RT-qPCR). Assays are detailed in **Table 1**. The prediction of the melting temperature ™ was performed *in silico* using the software uMELT Quartz (https://dna-utah.org/umelt/quartz/) (9). To establish the SYBR RT-qPCR, different primer concentrations (for E gene) and annealing temperatures (58°C as described at the original protocol and 60°C for all genes) were tested (**Table 2**). Synthetic RNA fragments from SARS-CoV-2 target genes were used as positive controls and for the generation of the standard curves for all assays.

**Table 2.** Performance of primers of the intercalating dye-based RT-qPCR protocol for SARS-CoV-2 detection

| Gene | Primer<br>concentration | 58°C* |  | 60 °C* |  |
| --- | --- | --- | --- | --- | --- |
|  |  | Tm (°C) | LOD (copies) | Tm (°C) | LOD (copies) |
| E | Fwd, Rv: 200nM | - | - | 81.02 (±0.07) | 50 |
|  | Fwd, Rv: 400nM | 81.06 (±0.03) | 50 | 81.07 (±0.02) | 5 |
| RdRp | Fwd: 600nM<br>Rv: 800nM | 80.6 (±0.03) | 1000 | 80.6 (±0.06) | 100 |

Fwd: forward primer, Rev: reverse primer
\* Primer annealing temperature
Tm: Melting temperature
LOD: Limit of detection

## Results

To provide alternatives for molecular detection of SARS-CoV-2 infections during material shortages, we compared the following: ordinary cotton swab “Q-tips” available in supermarkets against medical swabs, a classical (phenol/guanidine-based) protocol for RNA extraction against the commercially available kit for viral RNA extraction, and a cost-saving intercalating dye-based one-step, reverse transcription quantitative real-time PCR (SYBR RT-qPCR) protocol against the gold standard hydrolysis probe-based one-step, reverse transcription quantitative real-time PCR (probe RT-qPCR) for detection of SARS-CoV-2. These alternative options could also be beneficial for SARS-CoV-2 diagnosis in laboratories in low-resource settings.

### Comparison of swab types and RNA extraction methods

Participants self-sampled themselves using two sets of three different swab types (2x Q-tips, 2x flocked swabs, 2x rayon swabs) and RNA was extracted from each swab type either by using the classical protocol or the commercial kit. Of each swab type, the same quantity of input material was used for respective RNA extraction. To evaluate the applicability of these alternative swabs as options for self-sampling, the housekeeping gene hRNAse P was quantified to serve as endogenous control by RT-qPCR. Of non-infected participants, Q-tips swabs plus classical RNA extraction was as efficient for RNA recovery as the ones extracted by kit, with a Ct of 29±0.7 and 29±1.7, respectively (**Figure 1A**). Of SARS-CoV-2 infected participants, RNA recovery with Q-tips plus classical extraction was more efficient than the kit-based RNA extraction (Ct: 25±1.4 vs 28±3.8) (**Figure 1B**). Overall, sampling with ordinary Q-tips was as good as with medical swabs with rather slightly lower Ct values (indicating more RNA recovery in the extraction). Likewise, the classical RNA extraction protocol is at least as good as the commercial kit, when the control gene hRNAse P was amplified. As expected, flocked swab samples resulted in higher Ct values reflecting less recovery of RNA material compared to Q-tips and rayon swab. Flocked swabs are immersed in saline solution after sampling and only a fraction (140 µl of 2000 µl) is used for RNA extraction according to the standard protocol. This finding was consistent for sampling of uninfected as well as for SARS-CoV-2 positive individuals (**Figure 1A and 1B)**.

**Figure 1.**
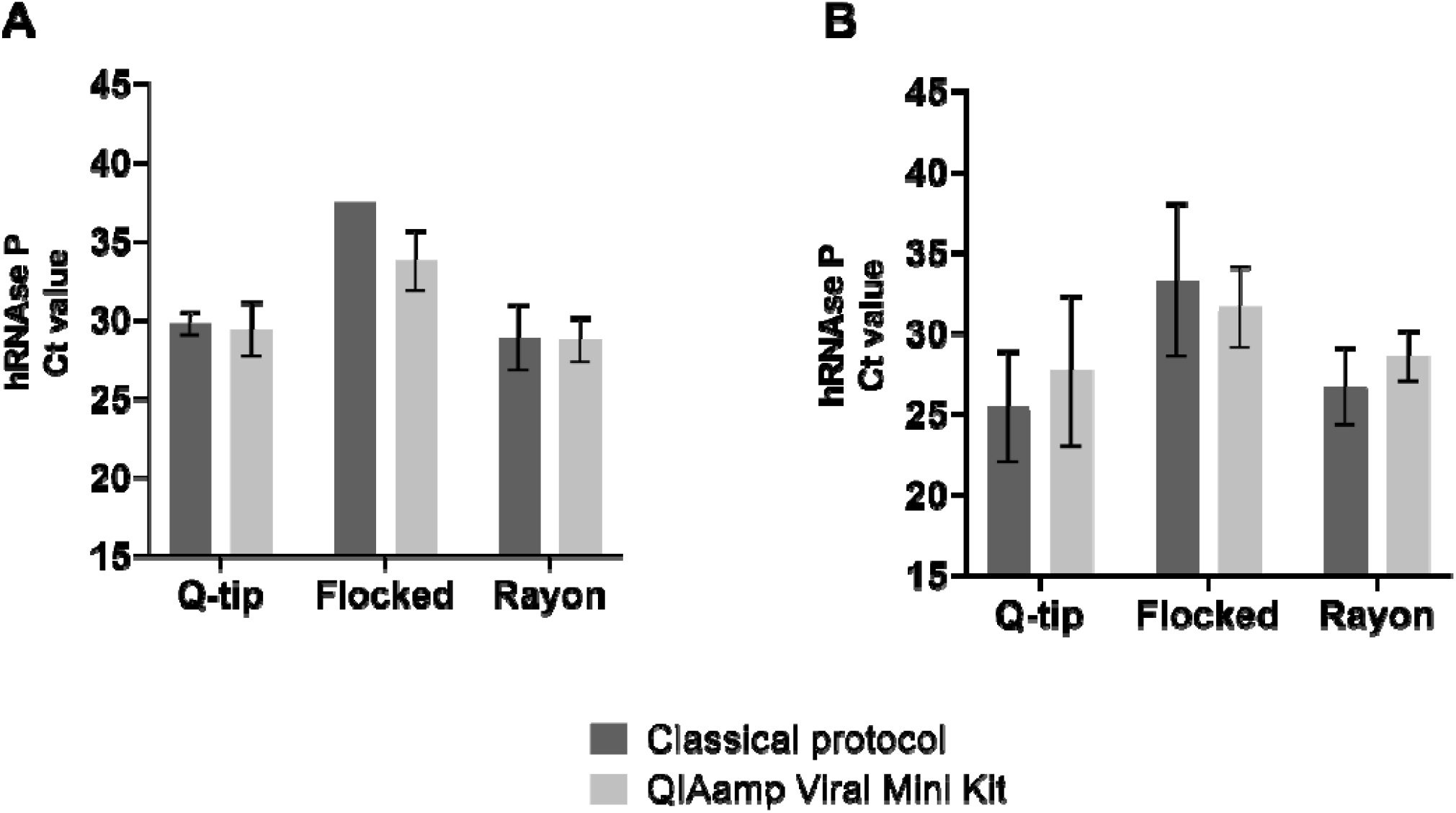
Comparison of swab types and RNA extraction methodologies for RNA extraction efficiency. Q-tips, flocked swabs, and rayon swabs were used, and RNA was extracted either by a classical protocol or the commercial kit. Ct values for hRNAse P amplification of oropharyngeal samples from **A**. non-infected participants (n=6); and **B**. SARS-CoV-2-infected participants (n=7).

### Establishment of an intercalating dye-based RT-qPCR for SARS-COV-2 detection

Next, we simplified the available SARS-CoV-2 probe RT-qPCR protocol towards an intercalating dye-based RT-qPCR (SYBR RT-qPCR) plus melting curve analysis as an alternative to the costly probe RT-qPCR assay. This was done first to detect the control gene hRNAse P and then for the SARS-CoV-2 genes E and RdRp. Performance of the E gene assay was more stable and more sensitive at a primer concentration of 400nM and the annealing temperature set to 60°C than primers at 200mM and/or at 58°C temperature as in the original protocol (**Table 2**). The RdRp gene assay also showed a better performance at 60°C (**Table 2**). *In silico* melting curve analysis predicted a melting temperature ™ of 85±0.43°C for hRNAse P, 81±0.43°C for E gene, and 80.5±0.43°C for RdRp. The hRNAse P, E, and RdRp genes assays presented a specific melting peak at 84.5 °C, 80.6 °C, and 81.1 °C, respectively. A few negative samples showed a unique unspecific melting peak at 75.9°C in the E gene assay that could be differentiated from the specific one.

When comparing the SYBR RT-qPCR to probe RT-qPCR, hRNAse P (LOD: 10 copies for both) and E gene (LOD: 5 copies for both) assays showed similar performance, while the RdRp gene assay using SYBR RT-qPCR was less sensitive compared to the probe RT-qPCR (LOD 100 vs. 10 copies) (**Figure 2**).

**Figure 2.**
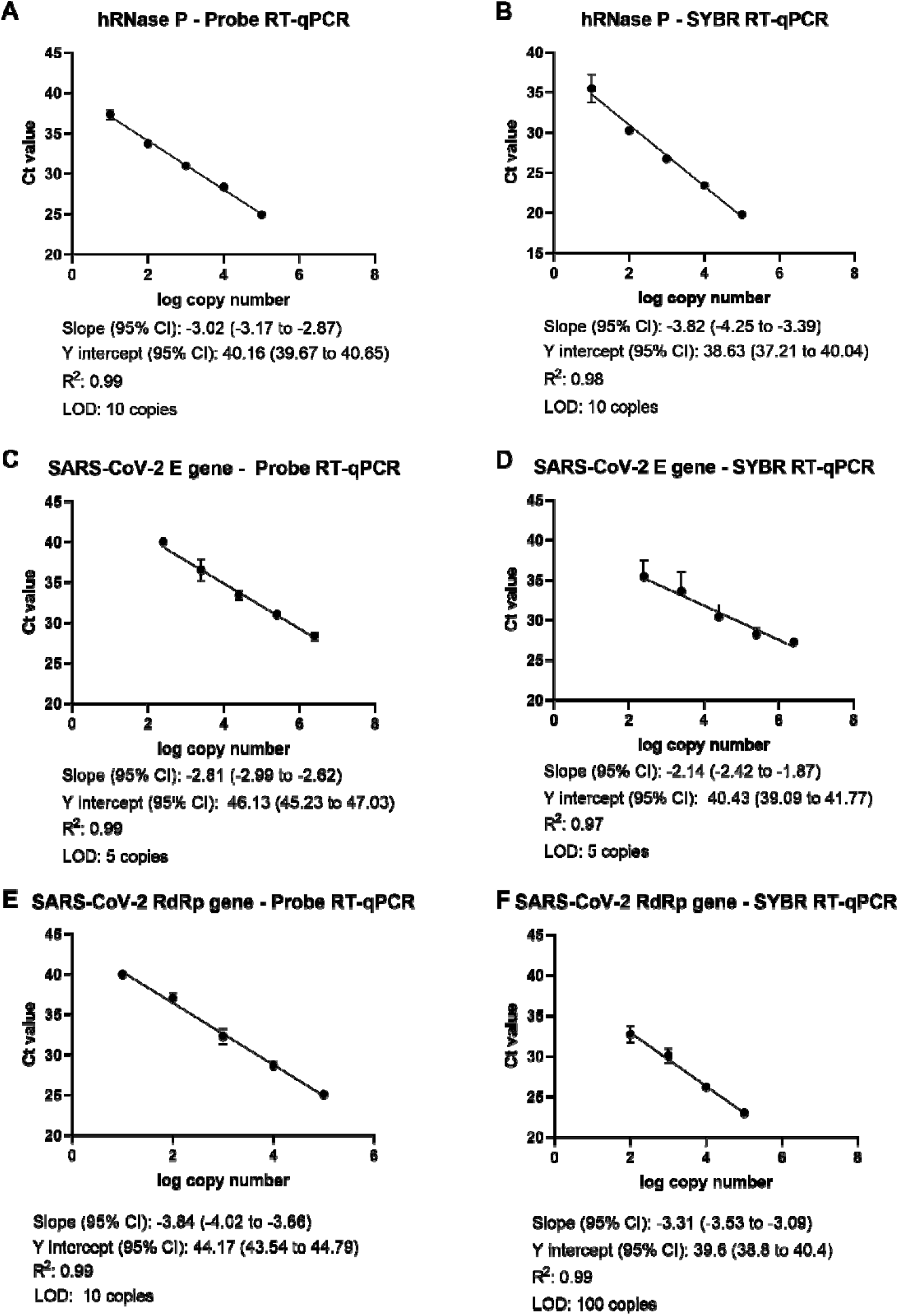
Comparing standard curves generated with SYBR green-based RT-qPCRs to hydrolysis probed-based RT-qPCR of the targeted genes

### Hydrolysis probe-based RT-qPCR assay

All target genes were consistently detected from swabs of all types (**Figure 3**). Considering the type of swab and the number of target gene copies, RNA recovered from Q-tips was similarly high as from rayon swabs (Q-tips: hRNAse P: 4.2±1.1 log copies, E: 5.1±1.1, and RdRp: 2.7±0.4 versus rayon swab: hRNAse P: 4.2±0.5 log copies, E: 5.2±0.7, and RdRp: 2.9±0.6). In contrast, flocked swab (hRNAse P: 3±0.9 log copies, E: 4.5±0.6, and RdRp: 2.1±0.8) was less suitable for cellular sampling and RNA yield (**Figure 3**).

**Figure 3.**
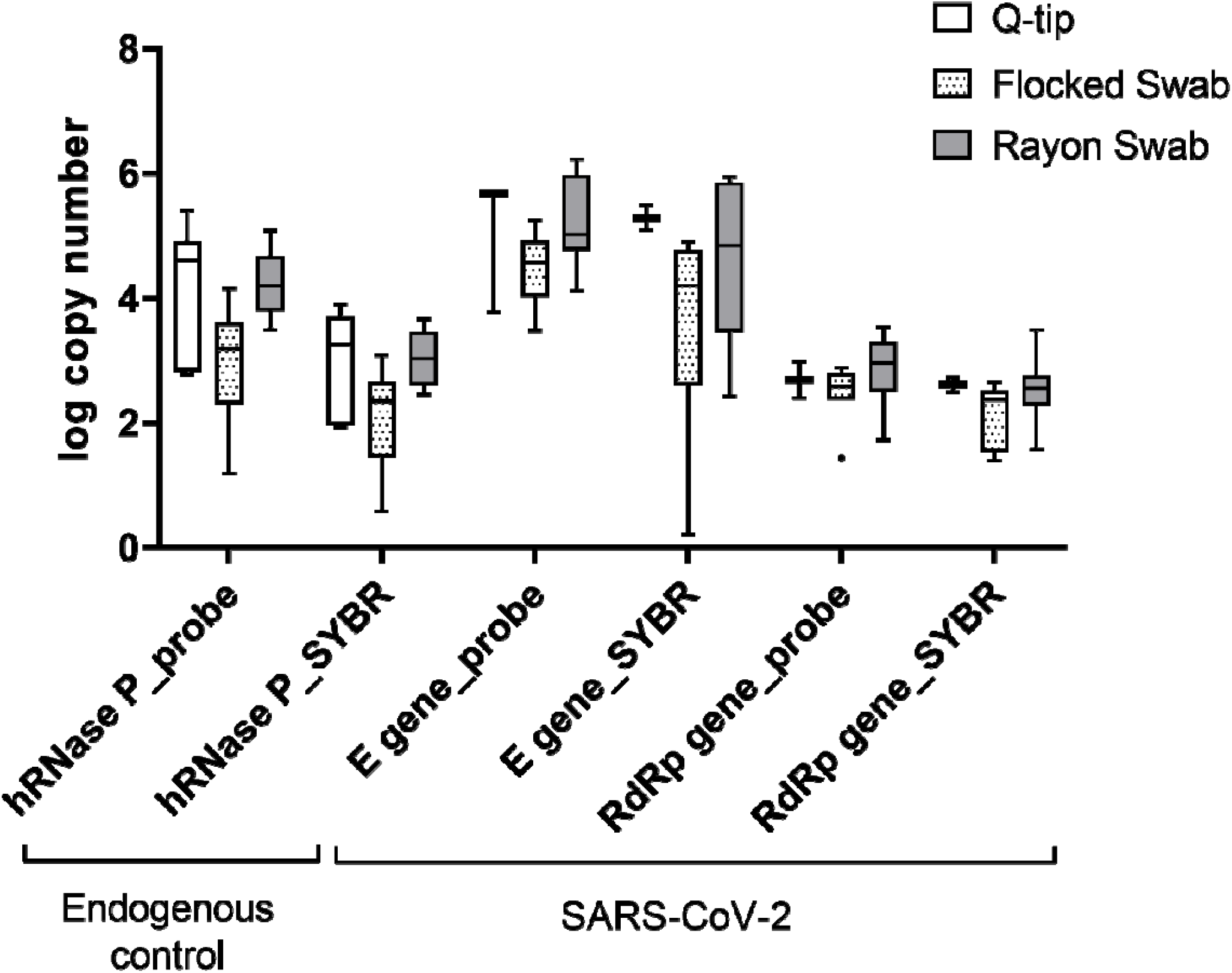
Comparison of the performance of the SYBR RT-qPCR and hydrolysis-probe based RT-qPCR methods in detecting the target genes in different swab types of non-infected (n = 6) and of SARS-CoV-2 infected participants (n= 7). RNA was extracted with the classical protocol or the commercial kit.

### SYBR RT-qPCR assay for SARS-CoV-2 detection as an alternative to probe RT-qPCR

Paired samples tested with both intercalating dye-based and probe-based assays showed a mean variance in the Ct values of 0.1 cycle for hRNAse P, 0.4 cycle for E gene, and 2.2 cycles for RdRp. The RdRp gene assay using intercalating dye presented earlier Ct values when compared to the probe-based. The intercalating dye-based assay failed in detecting one sample for both E gene (probe-based Ct value: 32.8) and RdRp gene (probe-based Ct value: 31.6) assays. Despite those failures, both intercalating assays could detect even later Ct values of other samples (the latest Ct value detected in field samples was 35.22 for E gene and 34.97 for the RdRp gene). Interestingly, both samples were extracted using the classical method. Regarding the log of gene copies compared to hydrolysis probe-based assays, the hRNAse P intercalating dye-based assay presented lower values when using rayon swab and Q-tips (R: 3.0±0.4 vs. 4.2±0.5 and Qt: 3.0±0.8 vs. 4.2±1.1) and equivalent results when using flocked swab+saline (2.1±0.8 vs. 2.1±0.5); E gene intercalating dye-based assay presented lower values when using rayon swab and flocked swab+saline (R: 4.6±1.3 vs. 5.2±0.7 and FS: 3.6±1.6 vs. 4.5±0.6) and equivalent results when using Q-tips (5.3±0.3 vs. 5.1±0.3); and RdRp gene assays presented consistent results among the different techniques utilized (R: 2.5±0.6 vs. 2.9±0.6, Qt: 2.6±0.2 vs. 2.7±0.4, and FS: 2.1±0.5 vs. 2.1±0.8) (**Figure 1**).

## Discussion

The current pandemic due to the SARS-CoV-2 rapid spread demands more testing and there is an urgent need for alternative but reliable materials for diagnosis. With a continuous global public health emergency state, it is predicted that several million people will become infected in the upcoming months of 2020/2021 (10, 11). Thus, reliable and accessible materials, reagents, and methods for diagnosis of SARS-CoV-2 infections are of paramount importance. Here, we compared the performance of ordinary Q-tips commonly available at supermarkets against medical flocked swabs and rayon swabs, a classical protocol for RNA extraction against a commercial kit and established a cost-efficient intercalating dye-based RT-qPCR for detection of SARS-CoV-2. This approach is not only urgently needed, it is also helpful to regions which suffer from limited financial resources. Overall, the performance of self-sampled oropharyngeal Q-tips, the phenol/guanidine-based (classical) protocol for RNA extraction, and the intercalating dye-based RT-qPCR for detection of SARS-CoV-2 genetic material was - individually as well as in combination – as reliable and sensitive as their respective comparators recommended by the authorities.

Here we also report the convenience and feasibility of swab self-sampling, the SARS-CoV-2 infected participants received kits with swabs, sampled themselves, and called the study team to collect them, reducing the SARS-CoV-2 spread and the study team exposure. In addition to this, in a pandemic scenario, self-sampling can also improve the mass screening participation, reduce the medical staff workload, and keep the clinics, hospitals, and even emergency rooms dedicated to the ones really in need of medical care. Also, in places with limited access to medical care, self-sampling with Q-tips swabs (available at the supermarket) that can be further collected may be an interesting option.

In all RT-qPCR assays either detecting hRNase P, which served as an endogenous control for sample integrity and proper RNA extraction, Ct values obtained for samples taken by Q-tips were similar to those obtained by rayon swab, while samples taken by flocked swab showed fewer copies. As recommended, flocked swabs are stored in 2 ml of saline after sampling and only 1/14 of the volume is used in the subsequent RT-qPCR assay. The reduced sensitivity is an obvious consequence of the reduced fraction tested.

The classical procedure for RNA extraction, done before the introduction of commercial kits became widely available, requires multiple handling steps including cell lyses using phenol/guanidine (or a commercially available solution such as QIAzol used here), phase separation with chloroform and RNA precipitation with isopropanol. This resulted into yields and qualities of extracted RNA that did not differ in Ct outputs of RT-qPCR amplification compared to RNA extracted with commercial kits with simplified and standardized pipetting steps. Interestingly, despite the fact that the used commercial kit is designed to isolate mainly viral RNA (QIAamp Viral RNA Mini Handbook), this did not result into lower Ct values of SARS-CoV-2 RT-qPCR and thus more starting copies compared to the classical protocol which extracts human and viral RNAs.

In contrast to hydrolysis probe-based RT-qPCR, intercalating dye-based assays do not depend on modified oligonucleotides where special manufacturing and supply can be critical in the current times. Intercalating dye-based RT-qPCR protocols have occasionally been used for SARS-CoV-2 diagnosis. Here, we expanded the available Charité protocol (8) towards a SYBR one-step RT-qPCR protocol for SARS-CoV-2 detection. *In silico* analysis of the primers and melting temperature peaks showed specificity and feasibility. The assay performance was sensitive and robust after increasing the annealing temperature from 58°C to 60°C compared to the probe-based protocol. The intercalating dye-based assays for hRNAse P and E genes were as sensitive as the hydrolysis probe-based assays, but the RdRp gene showed ten times less sensitivity. Dorlass and collaborators (12) also reported good performance of E gene primers published by Corman et al. (8) using intercalating dye-based assays in nasopharyngeal swabs collected in hospitals when compared to probe-based assay. Only one SARS-CoV-2 RT-qPCR positive individual who was positive for RNAse P by SYBR RT-qPCR was not positive for the E and RdRp genes when material from Q-tips plus classical RNA extraction was tested. In addition, one sample was not positive for the RdRp SYBR RT-qPCR assay despite the other two assays were successful. Thus, the intercalating dye-based RT-qPCR is a promising alternative for SARS-CoV-2 molecular diagnosis.

In pandemic scenarios, a shortage of recommended materials for molecular diagnostics can limit the identification of infected individuals which may have a negative impact on the efficiency of isolation measures and contributes to the spread of the virus. Moreover, the costs of the recommended materials often cannot be afforded by low-resource settings. Here, we showed that Q-tips available at very low costs in local supermarkets can be used for oropharyngeal self-sampling, RNA can be extracted with a classical (phenol-guanidine-based) protocol, and intercalating dye-based RT-qPCR is a valuable and sensitive diagnostic test reliably detecting SARS-CoV-2 infected individuals.

## Data Availability

On request the data can be shared.

## Acknowledgments

All authors are grateful to the participants. This publication was supported by the European Virus Archive Global (EVA-GLOBAL) project that has received funding from the European Union’s Horizon 2020 research and innovation program under grant agreement No 871029.

## Notes

### Competing Interest Statement

The authors have declared no competing interest.

### Clinical Trial

This is not a clinical trial

### Funding Statement

No external funding was received.

### Author Declarations

The study was approved by the Ethics Committee of the Universitaetsklinikum Tuebingen/Germany (Ref. number 20/231/B01).

